# Do Weather Temperature and Median-age affect COVID-19 Transmission?

**DOI:** 10.1101/2020.04.16.20067355

**Authors:** Aly Zein Elabdeen Kassem

## Abstract

It was observed that the coldest countries and the eldest in terms of median-age were most distressed by COVID-19 pandemic, while the warmest countries and that have younger-aged population were the least affected. Therefore, this study utilized the non-linear least squares method to estimate the impact of weather temperatures and median age on COVID-19 cases per million in thirty-nine countries divided into two groups. The first group composed of twenty-four countries that announced the first COVID-19 case in January 2020, while the second group contains fifteen countries that witnessed the pandemic for the first time in February of the same year. The study revealed some major findings, which are: COVID-19 cases per million were not significantly affected by weather temperature or the median age in “January-group” countries (after 72.67 days on average), while COVID-19 cases per million increased significantly by decreasing temperatures, and increasing the median age in case of “February-group” countries (after an average of 44.80 days). This means that weather temperature and median age may influence the transmission rates of COVID-19 in its early stages, while weather temperature or median age no longer have effects in the advanced stages of the pandemic.

## 1. Introduction

There is no doubt that the containment of COVID-19 pandemic caused by the emerging coronavirus (SARS-CoV-2) is currently the first concern worldwide. Since the characteristics of this pandemic that started in Wuhan, China in late 2019, made it difficult even by the most advanced health systems to control over it. This pandemic, which has spread to most countries of the world due its extremely high transmission rate of 2-2.5 (WHO, 2020-1), not only more targets the elder age categories, but it may also target specific places in the world, but rather specific areas in the country itself.

The number of COVID-19 cases globally as of April 10, 2020, at 14:27 GMT, was 1,625,213 cases, of which, 22% recovered, 5.95% died, and 71.47% were still active. According to official statistics, China, the source of the pandemic, managed to close 94.56% of cases with recovery, 4.07% with deaths (two cases per one million), and only 1.36% of cases were still active. While the USA topped the most affected countries’ list, with a number of cases amounted to 470,175, represented 28.93% of global cases, of which 90.9% cases were still active. Furthermore, the combined COVID-19 cases in Spain, Italy, Germany and France accounted for 33.09% of global cases as of study time (WORLDMETERS, 2020).

This paper investigates the impact of weather temperatures and median age individually on the number of COVID-19 cases per million.

## 2. Literature

### 2.1. Effect of temperature on COVID-19 transmission

A number of studies (Wang, Tang, Feng, & Lv, April 3, 2020; Sajadi & et. al., 2020; Chan, Peiris, Poon, & Seto, 2011; Casanova, Jeon, Rutala, Weber, & Sobsy, 2010; Altamimi & Ahmd, 2019) have proven that temperature and high relative humidity reduce the transmission rate of coronaviruses.

(Gundy & Gebra, 2009) have demonstrated that the activity of SARS virus in water is highly dependent on temperature, as the virus is rapidly inactivated in water at a temperature of 23°C for ten days compared to more than 100 days at a temperature of 4°C. In 2010 (Casanova, Jeon, Rutala, Weber, & Sobsy, 2010) recorded that SARS Cov was inactivated faster at 20°C compared to 4°C at all levels of relative humidity (above 20%), and that virus was inactivated faster at 40°C than 20°C. (Chan, Peiris, Poon, & Seto, 2011) explained in 2011 that SARS Cov virus retain its vitality for more than five days on surfaces at a temperature of 22-25°C and relative humidity between 40-50 %, But the virus soon loses its vitality at higher temperatures and relative humidity (such as 38°C, and relative humidity above 95%).

Although, research is still being carried out on the thermal tolerance of SARS-CoV-2, (Wang, Tang, Feng, & Lv, 2020) concluded that a temperature increase of 1°C and a rise in relative humidity by one degree would reduce the effective daily reproductive number of SARS-CoV-2 by 0.0225 and 0.0158, respectively. In the same vein, Sajadi & et. al. (2020) concluded that countries that witnessed a significant transmission of COVID-19 are roughly distributed along the 30-50 N’ corridor, which are countries with consistently climate patterns with temperatures ranging from 5-11°C, combined with low specific (3-6 g / kg) and absolute humidity (4-7 g / kg).

### 2.2. Age categories most affected by COVID-19

Several reports and studies have shown that elder age categories are more susceptible to COVID-19 ((WHO), 2020-1; Wei & Lessler, 2020); Ropert Koch Institut, 2020 (WHO), 2020-2; Kassem, 2020).

## 3. Material and Methods

To achieve the study’s objectives, thirty-nine countries were chosen and divided into two groups, twenty-four of which were declared the first COVID-19 case in January 2020, namely: the USA, Italy, Spain, China, Germany, France, UK, South Korea, Australia, Sweden, Malaysia, Japan, Russia, Philippines, Thailand, Finland, Singapore, Hong Kong, Taiwan, Vietnam, Sri Lanka, Cambodia, Macau and Nepal. While fifteen countries witnessed COVID-19 pandemic for the first time on February at the same year, which were: Iran, Switzerland, Belgium, Netherlands, Austria, Brazil, Israel, Norway, Ireland, Czech Republic, Denmark, Argentina, Egypt, Iraq and Lebanon.

Afterward, the temperature average in the most affected cities within the study’s two groups from the day of declaring the first case to April 5, 2020 were calculated (as shown in tables 3 and 6). Utilizing non-linear least squares method (Hansen, 2000), the relationships between cases of COVID-19 per million as a dependent variable for each of the study’s two groups, and the average temperatures in the most affected cities and median age as independent variables were estimated individually.

## 4. Description of the study Sample

### 4.1. Countries that reported the first case of COVID-19 in January 2020

Table (1) shows the most affected cities by COVID-19 in the twenty-four countries that witnessed the pandemic for the first time in January 2020. It is noteworthy that COVID-19 cases are mostly concentrated in the central and northern areas in these countries that represent capitals, densely populated cities, economic and financial centers, especially in developed countries. For instance, California, the USA’s economic capital, was the most affected in the far northeast. Likewise, the province of Lampodria in northern Italy, the responsible for 40% of industrial production^2^ was the most effected Italian area. Furthermore, COVID-19 has swept through Madrid, the Spanish capital, and the most important financial and economic center^3^. As well, the pandemic was concentrated in Wuhan, the transportation and industry hub in central China^4^, and the capital of China’s Hubei Province, the source of the SARS-CoV-2 virus. In Germany, the province of Bavaria, the second largest German province in terms of population, the producer of 18% of the gross German domestic product^5^ was the most affected. As for France, the pandemic targeted Ile-de-France region in north-central France, which is the richest region in Europe, and the most important French and European region in terms of research, development and innovation. In like manner, the pandemic was concentrated in London, the British capital, As well in Diego, the third largest city in South Korea, besides Stockholm, the Swedish capital, and in capitals of the remain countries in this group.

**Table 1:** Total cases and most effected cities in 24 countries that first experienced COVID-19 in January 2020

| Countries | 1st case | Days till 5<sup>th</sup> of April | Total cases | Cases per 1M | Most effected cites |
| --- | --- | --- | --- | --- | --- |
| USA | 20/01/2020 | 76 | 311,637 | 941 | New York |
| Spain | 30/01/2020 | 66 | 126,168 | 2699 | Madrid |
| Italy | 29/01/2020 | 67 | 124,632 | 2061 | Lombardy |
| Germany | 26/01/2020 | 70 | 96,092 | 1147 | Munich, Bavaria |
| France | 23/01/2020 | 73 | 89,953 | 1378 | Il de France; Paris |
| China | 10/01/2020 | 86 | 81,669 | 57 | Wuhan (Hubei) |
| UK | 30/01/2020 | 66 | 41,903 | 617 | London |
| S. Korea | 19/01/2020 | 77 | 10,237 | 200 | Daegu Metropolitan City |
| Sweden | 30/01/2020 | 66 | 6,443 | 638 | Stockholm |
| Australia | 24/01/2020 | 72 | 5,635 | 221 | New south wales; Sydney |
| Russia | 30/01/2020 | 66 | 4,731 | 32 | Mosco |
| Malaysia | 24/01/2020 | 72 | 3,483 | 108 | Selangor, Shah Alam |
| Japan | 14/01/2020 | 82 | 3,139 | 25 | Kanto; Tokyo |
| Philippines | 29/01/2020 | 67 | 3,094 | 28 | Manila |
| Thailand | 12/01/2020 | 84 | 2,169 | 31 | Bangkok |
| Finland | 28/01/2020 | 68 | 1,927 | 348 | Uusimaa |
| Singapore | 22/01/2020 | 74 | 1,189 | 203 | Singapore |
| Hong-Kong | 22/01/2020 | 74 | 862 | 115 | Hong Kong capital city-state |
| Taiwan | 20/01/2020 | 76 | 363 | 15 | Taipei |
| Vietnam | 22/01/2020 | 74 | 240 | 2 | Hanoi |
| Sri Lanka | 26/01/2020 | 70 | 166 | 8 | Sri Jayawardenepura Kotte |
| Cambodia | 26/01/2020 | 70 | 114 | 7 | Phnom Penh |
| Macao | 21/01/2020 | 75 | 44 | 68 | Municipality of Macau |
| Nepal | 23/01/2020 | 73 | 9 | 0.3 | Kathmandu |

Sources: https://www.worldometers.info/ (Last accessed; 5<sup>th</sup> April, 2020, 06:49 GMT).

Tables (1) and (2) show that, despite the “January-group” countries experienced the pandemic at close times, however, these countries varied in the total number of COVID-19 cases, which averaged about 38 thousand, with a minimum of nine only in Nepal, and a maximum of about 312 thousand in the USA in the last update of this study’s data on April 5, 2020 at 6:49 GMT. Further, the study countries also varied in infection and death rates, and the number of virus detection tests per million citizens. Tables (2) and (3) show that the “January-group” countries differed in average weather temperatures during the pandemic days until the fifth of April 2020. The average temperatures in these countries reached about 14.68°C with a minimum of −1.83 in Finland, and a maximum of 30.50 in Thailand.

**Table 2:**
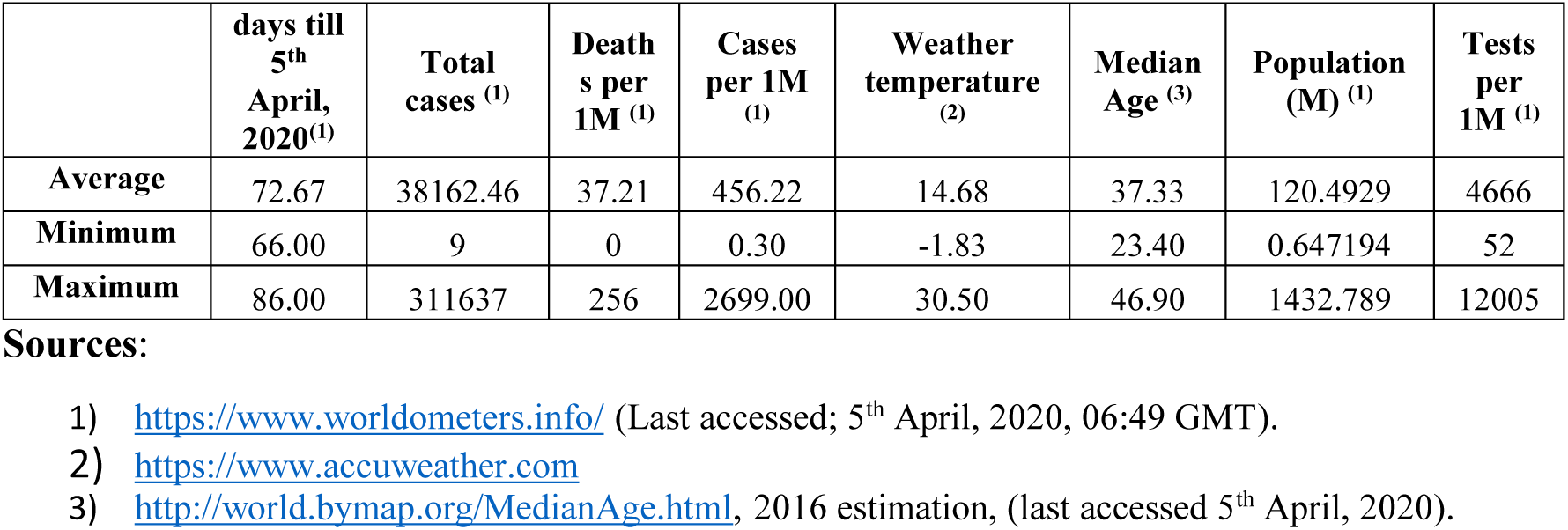
COVID-19 spread indicators, Weather temperatures and median age in 24 countries that first experienced COVID-19 in January 2020

|  | days till 5<sup>th</sup> April, 2020<sup>(1)</sup> | Total cases <sup>(1)</sup> | Deaths per 1M <sup>(1)</sup> | Cases per 1M <sup>(1)</sup> | Weather temperature <sup>(2)</sup> | Median Age <sup>(3)</sup> | Population (M) <sup>(1)</sup> | Tests per 1M <sup>(1)</sup> |
| --- | --- | --- | --- | --- | --- | --- | --- | --- |
| Average | 72.67 | 38162.46 | 37.21 | 456.22 | 14.68 | 37.33 | 120.4929 | 4666 |
| Minimum | 66.00 | 9 | 0 | 0.30 | -1.83 | 23.40 | 0.647194 | 52 |
| Maximum | 86.00 | 311637 | 256 | 2699.00 | 30.50 | 46.90 | 1432.789 | 12005 |

Sources:
- 1) https://www.worldometers.info/ (Last accessed; 5<sup>th</sup> April, 2020, 06:49 GMT). - 2) https://www.accuweather.com - 3) http://world.bymap.org/MedianAge.html, 2016 estimation, (last accessed 5<sup>th</sup> April, 2020).

**Table 3:** Average temperature and median age in 24 countries that first experienced COVID-19 in January 2020

| Most effected cites | Temperature at <sup>(1)</sup> |  |  |  |  |  |  | Median Age<sup>(4)</sup> |
| --- | --- | --- | --- | --- | --- | --- | --- | --- |
|  | First case-day |  | February <sup>(2)</sup> |  | April 5, 2020 |  | Monthly average temp. <sup>(3)</sup> |  |
|  | High | Low | High | Low | High | Low |  |  |
| New York, USA | -1 | -7 | 2 | -3 | 11 | 5 | 1.8 | 37.9 |
| Madrid, Spain | 11 | 8 | 14 | 8 | 16 | 5 | 10.3 | 42.3 |
| Lombardy, Italy | 8 | -2 | 13 | 0 | 17 | 4 | 6.7 | 45.1 |
| Munich, Bavaria, Germany | 9 | -3 | 13 | 6 | 16 | 0 | 6.8 | 46.8 |
| Il de France; Paris, France | 4 | -1 | 13 | 9 | 9 | 2 | 6.0 | 41.2 |
| Wuhan (Hubei), China | 8 | 0 | 13 | 4 | 13 | 6 | 7.3 | 37.1 |
| London, England | 12 | 5 | 10 | 3 | 8 | 2 | 6.7 | 40.5 |
| Daegu Metropolitan City, S. Korea | 9 | -4 | 8 | -2 | 18 | 4 | 5.5 | 41.2 |
| Stockholm, Sweden | 3 | -2 | 5 | -4 | 2 | -4 | 0.0 | 41.2 |
| New south wales; Sydney, Australia | 32 | 22 | 29 | 19 | 26 | 20 | 24.7 | 36.8 |
| Mosco, Russia | 0 | -1 | 1 | -3 | 2 | -4 | -0.8 | 39.3 |
| Selangor, Shah Alam, Malaysia | 36 | 25 | 35 | 25 | 35 | 26 | 30.3 | 28.2 |
| Kanto; Tokyo, Japan | 14 | 5 | 11 | 5 | 10 | 8 | 8.8 | 46.9 |
| Manila, Philippines | 30 | 24 | 32 | 25 | 34 | 27 | 28.7 | 23.4 |
| Bangkok, Thailand | 34 | 26 | 35 | 25 | 36 | 27 | 30.5 | 37.2 |
| Uusimaa, Finland | 3 | 0 | 0 | -9 | 2 | -7 | -1.8 | 42.4 |
| Singapore, Singapore | 32 | 26 | 31 | 26 | 35 | 27 | 29.5 | 34.3 |
| Hong Kong capital city-state | 19 | 14 | 26 | 17 | 21 | 20 | 19.5 | 44.0 |
| Taipei, Taiwan | 20 | 14 | 20 | 15 | 24 | 19 | 18.7 | 40.2 |
| Hanoi, Vietnam | 24 | 18 | 24 | 17 | 25 | 17 | 20.8 | 30.1 |
| Sri Jayawardenepura Kotte, Sri Lanka | 32 | 24 | 33 | 26 | 34 | 26 | 29.2 | 32.5 |
| Phnom Penh, Cambodia | 34 | 23 | 35 | 23 | 38 | 27 | 30.0 | 24.9 |
| Municipality of Macau, Macao | 22 | 15 | 21 | 14 | 20 | 18 | 18.3 | 38.7 |
| Kathmandu, Nepal | 18 | 2 | 23 | 7 | 29 | 11 | 15.0 | 23.6 |

**Sources:**
- 1) https://www.accuweather.com/en/es/madrid/308526/weather-forecast/308526 - 2) At the same first case-day, but in February - 3) The mean of the lower and upper temperatures in six observations from the first case-day in January until 5<sup>th</sup> April, 2020 - 4) http://world.bymap.org/MedianAge.html, 2016 estimation, (last accessed 5<sup>th</sup> April, 2020).

On the other hand, it is clear from Table (2) that the median age in the “January-group” countries has reached about 37.33 years, with a minimum of 23.40 years in Philippines, and a maximum of 46.90 years in Japan.

### 4.2. Countries that reported the first case of COVID-19 in January 2020

It is clear from Table (4) that, unlike the “holy city of Qom” in Iran, the COVID-19 pandemic was mainly concentrated in the capitals of countries that witnessed the pandemic in February 2020.

**Table 4:** Total cases and most effected cities in 15 countries that first experienced COVID-19 in February 2020

| **Countries** | **1st case** | **Days till 5<sup>th</sup> of April** | **Total cases** | **Cases per 1M** | **Most effected cites** |
| --- | --- | --- | --- | --- | --- |
| Iran | 18/02/2020 | 47 | 55,743 | 664 | Qom |
| Switzerland | 24/02/2020 | 41 | 20,505 | 2,369 | Bern |
| Belgium | 03/02/2020 | 62 | 18,431 | 1,590 | Brussel |
| Holland | 26/02/2020 | 39 | 16,627 | 970 | Amsterdam |
| Austria | 24/02/2020 | 41 | 11,781 | 1,308 | Vienna |
| Brazil | 24/02/2020 | 41 | 10,360 | 49 | Brasilia |
| Israel | 20/02/2020 | 45 | 8,018 | 926 | Tal Aviv |
| Norway | 25/02/2020 | 40 | 5,550 | 1,024 | Oslo |
| Ireland | 28/02/2020 | 37 | 4,604 | 932 | Dublin |
| Czech R. | 29/02/2020 | 36 | 4,475 | 418 | Prague |
| Denmark | 26/02/2020 | 39 | 4,077 | 704 | Copenhagen |
| Argentina | 02/02/2020 | 63 | 1,451 | 32 | Buenos Aires |
| Egypt | 13/02/2020 | 52 | 1,070 | 10 | Cairo |
| Iraq | 21/02/2020 | 44 | 878 | 22 | Baghdad |
| Lebanon | 20/02/2020 | 45 | 520 | 76 | Beirut |

**Sources:** https://www.worldometers.info/ (Last accessed; 5<sup>th</sup> April, 2020, 06:49 GMT).

It appears from Tables (4), (5) that despite the participation of all countries in this group at the beginning of the pandemic in February 2020, however, these countries also varied in the number of cases of COVID-19, which averaged about 10.94 thousand cases, with a minimum of only 520 cases in the case of Lebanon, and about 55.74 thousand in Iran as a maximum on the fifth of April 2020 at 6:49 GMT. The countries of this group also varied in infection and death rates, and the number of COVID-19 detection tests per million citizens.

**Table 5:** COVID-19 spread indicators, Weather temperatures and median age in 15 countries that first experienced COVID-19 in February 2020

|  | days till<br>5<sup>th</sup> April,<br>2020<sup>(1)</sup> | Total<br>cases <sup>(1)</sup> | Deaths<br>per 1M<br><sup>(1)</sup> | Cases<br>per 1M<br><sup>(1)</sup> | Weather<br>temperat<br>ure <sup>(2)</sup> | Median<br>Age <sup>(3)</sup> | Population<br>(M) <sup>(1)</sup> | Tests<br>per<br>1M <sup>(1)</sup> |
| --- | --- | --- | --- | --- | --- | --- | --- | --- |
| **Average** | 44.80 | 10939.33 | 28.72 | 739.6 | 12.34 | 34.96 | 38.43 | 6409.7<br>3 |
| **Minimum** | 36 | 520 | 0.1 | 10 | 3.50 | 19.90 | 4.94 | 188 |
| **Maximum** | 63 | 55743 | 111 | 2369 | 24.50 | 43.80 | 211.43 | 19528 |

Sources:
- 1) https://www.worldometers.info/ (Last accessed; 5<sup>th</sup> April, 2020, 06:49 GMT). - 2) https://www.accuweather.com - 3) http://world.bymap.org/MedianAge.html, 2016 estimation, (last accessed 5<sup>th</sup> April, 2020).

Tables (5) and (6) show that the countries that witnessed the pandemic in February differed in the average weather temperatures during the pandemic days until the fifth of April 2020. The average temperatures in these countries reached about 12.34 degrees Celsius, with a minimum of 3.50 in Norway, and a maximum 24.50 in Argentina. On the other hand, Table (5) shows that the median age in the countries of this group reached 34.96 years, with a minimum of 19.90 years in Iraq, and a maximum of 43.80 years in Austria.

**Table 6:** Average temperature and median age in 15 countries that first experienced COVID-19 in February 2020

| Most effected cites | Temperature at <sup>(1)</sup> |  |  |  |  |  | Median Age<sup>(4)</sup><br>First case-day<br>High | Most effected<br>cites<br>February <sup>(2)</sup><br>Low |
| --- | --- | --- | --- | --- | --- | --- | --- | --- |
|  | First case-day |  | February <sup>(2)</sup> |  | April 5, 2020 |  |  |  |
|  | High | Low | High | Low | High | Low |  |  |
| Qom | 16 | 1 | 21 | 11 | 20 | 8 | 12.83 | 29.2 |
| Bern | 9 | 2 | 18 | 2 | 18 | -2 | 7.83 | 42.2 |
| Brussel | 11 | 6 | 8 | 2 | 22 | 5 | 9.00 | 41.1 |
| Amsterdam | 7 | 2 | 11 | 1 | 20 | 5 | 7.67 | 42.5 |
| Vienna | 10 | 4 | 8 | -3 | 17 | -1 | 5.83 | 43.8 |
| Brasilia | 26 | 21 | 25 | 20 | 31 | 22 | 24.17 | 31.6 |
| Tal Aviv | 14 | 8 | 8 | 5 | 28 | 12 | 12.50 | 29.7 |
| Oslo | 1 | -4 | 9 | 2 | 10 | 3 | 3.50 | 39.1 |
| Dublin | 13 | 3 | 9 | 4 | 15 | 6 | 8.33 | 36.4 |
| Prague | 8 | -1 | 8 | -1 | 16 | 5 | 5.83 | 41.7 |
| Copenhagen | 7 | 3 | 8 | 2 | 11 | 5 | 6.00 | 42.0 |
| Buenos Aires | 32 | 22 | 31 | 24 | 24 | 14 | 24.50 | 31.5 |
| Cairo | 24 | 13 | 18 | 13 | 29 | 15 | 18.67 | 23.8 |
| Baghdad | 23 | 7 | 22 | 8 | 30 | 18 | 18.00 | 19.9 |
| Beirut | 17 | 14 | 13 | 9 | 16 | 34 | 17.17 | 29.9 |

Sources:
- 1) https://www.accuweather.com/en/es/madrid/308526/weather-forecast/308526 - 2) At the same first case-day, but in February - 3) The mean of the lower and upper temperatures in six observations from the first case-day in January until 5<sup>th</sup> April, 2020 - 4) http://world.bymap.org/MedianAge.html, 2016 estimation, (last accessed 5<sup>th</sup> April, 2020).

## 5. Results

### 5.1. Relationship between COVID-19 cases per million and temperature in “January-group” countries

The exponential function was suggested as in equation (1) to represent the relationship between the number of COVID-19 cases per million as a proxy of transmission as a dependent variable (y), and the weather temperature as an independent variable (x).

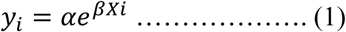

Where: y_i_: Number of cases of COVID-19 in country i.

α, β: The model parameters

x_i_: Average temperature in the country “i” since the beginning of the first case in January 2002, and until the fifth of April 2020.

Taking the natural logarithm of both sides of equation (1), the following equivalent equation can be obtained:

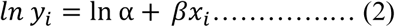

Where it was possible by converting to equation (2), obtaining a formula for a linear regression model, to which the error component ε can be added, to become as follows:

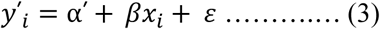

By applying the non-linear least square method using the mathematical formula as in equation (3), the relationship parameters between the total number of COVID-19 cases in the “January-group” countries, and weather temperatures were estimated as in table (7).

**Table 7:** Regression model between COVID-19 cases and weather temperature in countries that first experienced COVID-19 in January 2020

| Parameters | Values |
| --- | --- |
| No. of observations | 24 |
| Adjusted R Square | 0.0798 |
| F | 1.91 |
| Probability > F | 0.1810 |
| Intercept (Standard Error) | 5.4586 (1.6039) |
| P-value | 0.181 |
| Temperature (X) (Standard Error) | -0.12081 (0.0874) |
| P-value | 0.003 |

**Sources:** Tables (1) and (3), Using STATA 13.1.

The results in table (7) showed an inverse relationship between the number of cases of COVID-19 per million and temperatures in this group of countries, despite the lack of statistical significance for this relationship.

### 5.2. Relationship between COVID-19 cases per million and median age in “January-group” countries

Utilizing the same exponential formula previously described in equations (1, 2, and 3), it was possible to estimate the parameters of the relationship between COVID-19 cases per million and the median age as in Table (8). Where the results revealed a positive non-statistically significant relationship between COVID-19 cases per million and median age in “January-group” countries.

**Table 8:**
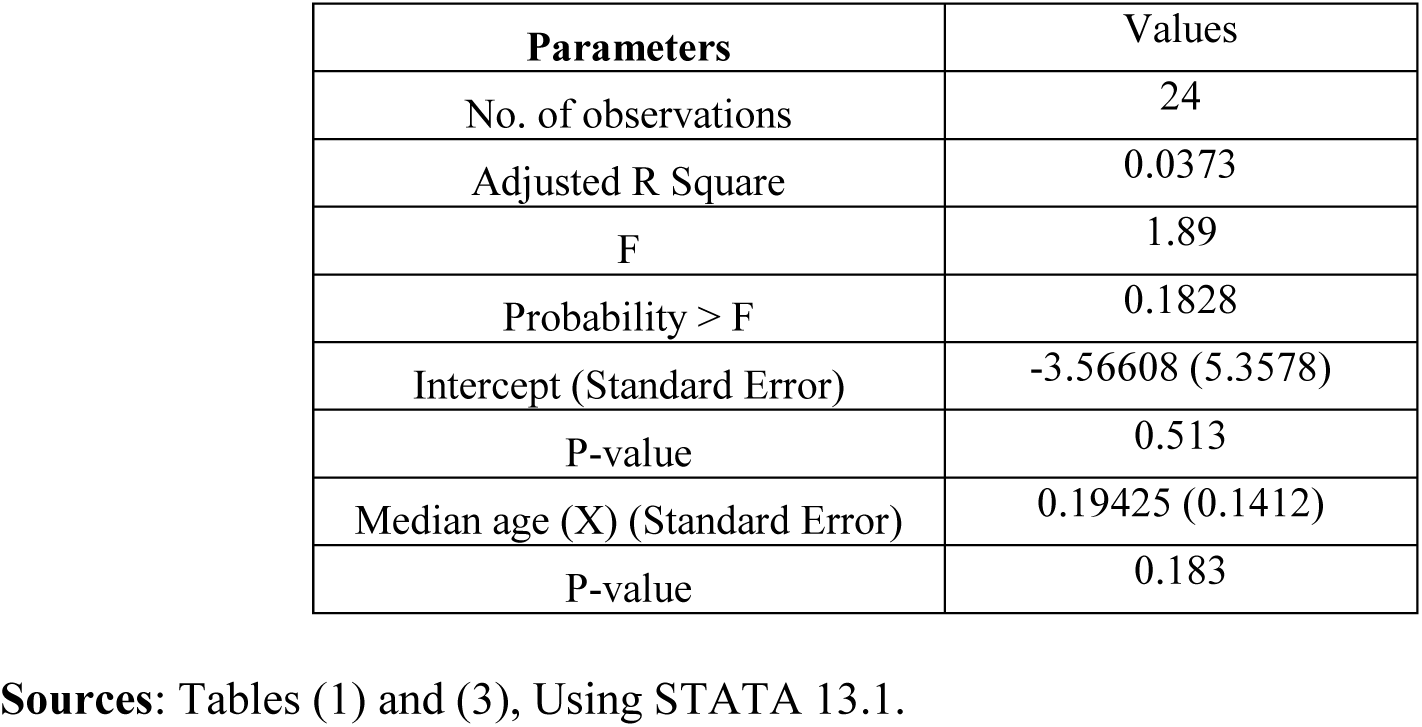
Regression model between COVID-19 cases and median age in 24 countries that first experienced COVID-19 in January 2020

| Parameters | Values |
| --- | --- |
| No. of observations | 24 |
| Adjusted R Square | 0.0373 |
| F | 1.89 |
| Probability > F | 0.1828 |
| Intercept (Standard Error) | -3.56608 (5.3578) |
| P-value | 0.513 |
| Median age (X) (Standard Error) | 0.19425 (0.1412) |
| P-value | 0.183 |

**Sources:** Tables (1) and (3), Using STATA 13.1.

### 5.3. Relationship between COVID-19 cases per million and temperature in “February-group” countries

As well, employing the same exponential formula previously described in equations (1, 2, and 3), the parameters of the relationship between COVID-19 cases per million and temperature was estimated as in Table (9). Where the results revealed a reverse statistically significant relationship between COVID-19 cases per million and temperature in “February-group” countries.

**Table 9:** Regression model between COVID-19 cases and temperature in 15 countries that first experienced COVID-19 in February 2020

| Parameters | Values |
| --- | --- |
| No. of observations | 15 |
| Adjusted R Square | 0.6599 |
| F | 18.17 |
| Probability > F | 0.0001 |
| Intercept (Standard Error) | 8.46565 (0.5812) |
| P-value | 0.000 |
| Median age (X) (Standard Error) | -0.22152 (0.0417) |
| P-value | 0.000 |

Sources: Tables (4) and (6), Using STATA 13.1.

Substitution by the model (2) parameters as in table (9):

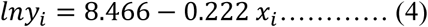

Applying e on both sides:

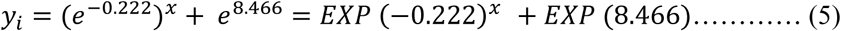

Equation (5) can be used for predicting the development of COVID-19 cases numbers in light of temperature as shown in Appendix (1).

### 5.4. Relationship between COVID-19 cases per million and temperature in “February-group” countries

The same manner, using the same exponential formula previously described in equations (1, 2, and 3), the relationship between COVID-19 cases per million and median age was estimated as in Table (10).

**Table 10:** Regression model between COVID-19 cases and temperature in 15 countries that first experienced COVID-19 in February 2020

| Parameters | Values |
| --- | --- |
| No. of observations | 15 |
| Adjusted R Square | 0.5969 |
| F | 21.73 |
| Probability > F | 0.0004 |
| Intercept (Standard Error) | -0.80334 (1.4320) |
| P-value | 0.584 |
| Median age (X) (Standard Error) | 0.18691 (0.0410) |
| P-value | 0.000 |

Sources: Tables (4) and (6), Using STATA 13.1.

Where the results revealed a positive statistically significant relationship between COVID-19 cases per million and median age in “February-group” countries.

Substitution by the model (2) parameters as in table (10):

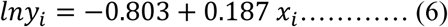

Applying e on both sides:

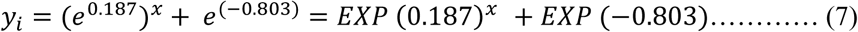

Furthermore, Equation (7) can be used for predicting the development of COVID-19 cases numbers in light of median age as shown in Appendix (2).

## 6. Discussion

Although the results of the study show that COVID-19 cases per million have increased in countries that have lower weather temperature, as well in countries have higher median age. However, this relationship is not absolute, but rather is related to the number of days since the first case appeared. Where the lower the time from the pandemic starting day, the higher the impact of temperature and median age on the transmission rates. In other words, temperature and median age affect COVID-19 transmission in its early stages, but when cases per million reach a critical mass after successive exponential increase, these two factors no longer have significant influence on the pandemic transmission.

Besides temperature and median age, there are other important factors that have worsened the situation in countries that were heavily invaded by COVID-19, such as the USA, Spain, and Italy, such as the delay in applying mitigation measures for instance. On the contrary to similar countries in terms of temperature at the earliest time of the pandemic i.e. China and South Korea or even Japan that have the higher median age among all the thirty-nine countries in this study, and managed to flatten the exponential growth curve of the pandemic by distinguished mechanisms of early mitigation measures they have applied and utilization of big data techniques in containing the pandemic from its primarily sources.

Referring to the prediction appendices numbers (1), (2), it turns out from table (11) that the number of COVID-19 cases per million in Iran, Switzerland, Belgium, and Israel were higher than its expected values, whether in terms of temperature or median age. As for Norway and Ireland, the registered cases per million were higher than expected in terms of median age, although they were lower than expected as regards to temperature. On the other hand, COVID-19 cases per million in Austria, Czech Republic, Denmark, Egypt and Lebanon were lower than expected in light of both temperature and median age. Almost in like manner, the pandemic cases per million in Netherlands, Brazil, and Argentina were lower than expected pertaining to median age, although these rates were slightly higher than expected as far as temperature. As for Iraq, COVID-19 cases per million were lower than expected in the light of temperature, although these rates were slightly higher than expected according to the median age.

**Table 11:** Observed and estimated COVID-19 cases per million* in terms of temperature and median age in the “February-group” countries

| Countries | Temperature | Median age | Observed COVID-19 cases per 1M | Estimated COVID-19 cases per 1M, in terms of |  |
| --- | --- | --- | --- | --- | --- |
|  |  |  |  | Temp. | M. age |
| Iran | 12.83 | 29.2 | 664 | 313 | 102 |
| Switzerland | 7.83 | 42.2 | 2,369 | 941 | 1154 |
| Belgium | 9.00 | 41.1 | 1,590 | 770 | 955 |
| Holland | 7.67 | 42.5 | 970 | 921 | 1163 |
| Austria | 5.83 | 43.8 | 1,308 | 1456 | 1661 |
| Brazil | 24.17 | 31.6 | 49 | 28 | 175 |
| Israel | 12.50 | 29.7 | 926 | 411 | 121 |
| Norway | 3.50 | 39.1 | 1,024 | 2044 | 658 |
| Ireland | 8.33 | 36.4 | 932 | 1001 | 378 |
| Czech R. | 5.83 | 41.7 | 418 | 1456 | 1141 |
| Denmark | 6.00 | 42 | 704 | 1498 | 1149 |
| Argentina | 24.50 | 31.5 | 32 | 29 | 174 |
| Egypt | 18.67 | 23.8 | 10 | 83 | 40 |
| Iraq | 18.00 | 19.9 | 22 | 104 | 19 |
| Lebanon | 17.17 | 29.9 | 76 | 131 | 122 |

**Sources:** Author, Tables (4, 6) and Tables (1, 2-Annex).
\* For April 5, 2020
Observed cases / million is less than estimated
Observed cases / million is slightly higher than estimated
Observed cases / million is higher than estimated

This study’s results assume that the data provided by countries to the international organizations are correct and accurate. But in case of assuming that the differences between the observed and expected COVID-19 cases in Table (11) are due to underestimation or underreporting, the number of expected total cases per million in some of these countries can be estimated as in table (12).

**Table 12:**
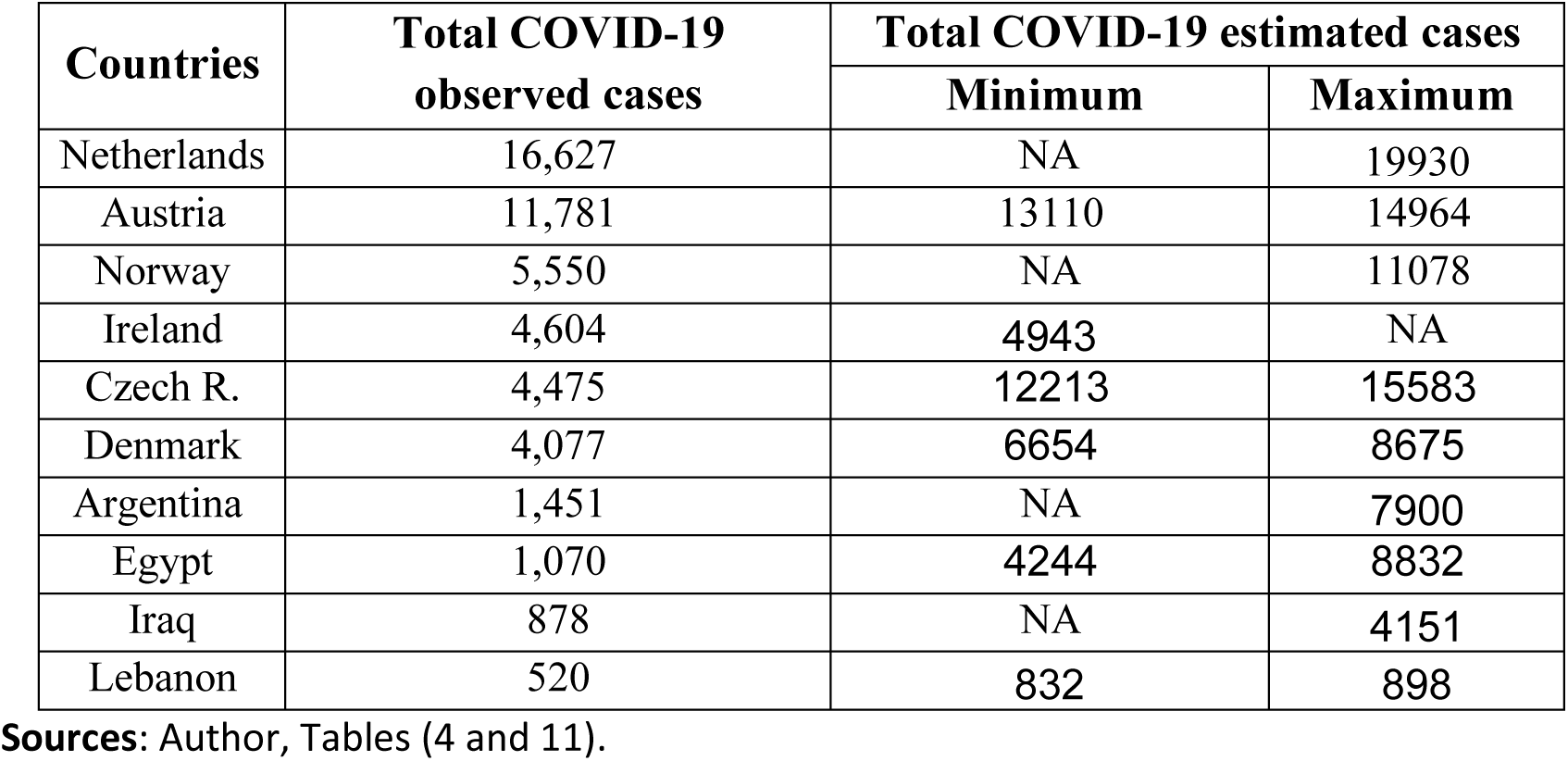
Observed and estimated total COVID-19 cases in some “February-group” countries on April 5, 2020

Generally, the results of this study may raise the question about the extent of the specialty of COVID-19, and its focus on geographical areas in which the major industrial countries concentrate, especially the economic and financial centers. It’s highly recommended that the relationship between COVID-19 cases per million and temperature and median age to be estimated at different time periods thereafter, in order to monitor the phenomenon at other time periods, either in later stages or more early than that observed in this study.

## 7. Conclusion

There are several factors that affect the increasingly transmission of COVID-19, among these factors are the decrease in weather temperature and the increase in the median age of the population. Although, temperature and median age may affect the rates of COVID-19 prevalence in its early stages, but when cases per million reach a critical mass after successive exponential increase, these two factors no longer have significant influence on the pandemic transmission.

## Data Availability

All data referred to in the manuscript are available

https://www.worldometers.info/coronavirus/

## Electronic sources

(ACCUWEATHER) https://www.accuweather.com

**BBC News (2020), Coronavirus:** Italian economy takes a body blow, **available at: https://www.bbc.com/news/business-51650974, (Accessed: 3 April 2002)**

(CITYPOPULATION) https://www.citypopulation.de

European Commission, **Internal Market, Industry, Entrepreneurship and SMEs**, Available at: https://ec.europa.eu/growth/tools-databases/regional-innovation-monitor/base-profile/ile-de-france, (Last accessed: 7 April, 2020)

European Commission, **Internal Market, Industry, Entrepreneurship and SMEs**, Available at: https://ec.europa.eu/growth/tools-databases/regional-innovation-monitor/base-profile/bavaria, (Last accessed: 7 April, 2020)

Eurostat, **First population estimates: EU population up to over 513 million on 1 January 2019 More death than birth**, (Brussels, Eurostat News release, 114/2019, 10 July 2019), available at: https://ec.europa.eu/eurostat/documents/2995521/9967985/3-10072019-BP-EN.pdf/e152399b-cb9e-4a42-a155-c5de6dfe25d1

Organization of Economic Cooperation and Development (OCED), **Lombardy, Italy**, (Paris, OCED, without year), available at: https://www.oecd.org/fr/sites/eduimhe/49008527.pdf

South China Morning Post (2020), **Why Wuhan is so important to China’s economy and the potential impact of coronavirus**, Available at: https://www.scmp.com/economy/china-economy/article/3047426/explained-why-wuhan-so-important-chinas-economy-and-potential, (Accessed: 7 April, 2020)

(WORLDMETERS) https://worldpopulationreview.com

(WORLDPOPULATIONREVIEW) https://worldpopulationreview.com/world-cities/madrid-population/

(WORLD.BYMAP), http://world.bymap.org/MedianAge.html

## Appendices

**Appendix (1):** Prediction table of COVID-19 cases per million and temperature after 44.80 days of COVID-19 pandemic*

| Temp. °C | Cases per 1M | Temp. °C | Cases per 1M | Temp. °C | Cases per 1M |
| --- | --- | --- | --- | --- | --- |
| -5 | 17223 | 13 | 317 | 31 | 6 |
| -4 | 13794 | 14 | 254 | 32 | 5 |
| -3 | 11048 | 15 | 203 | 33 | 4 |
| -2 | 8848 | 16 | 163 | 34 | 3 |
| -1 | 7087 | 17 | 130 | 35 | 2 |
| 0 | 5676 | 18 | 104 | 36 | 2 |
| 1 | 4546 | 19 | 84 | 37 | 2 |
| 2 | 3641 | 20 | 67 | 38 | 1 |
| 3 | 2916 | 21 | 54 | 39 | 1 |
| 4 | 2336 | 22 | 43 | 40 | 1 |
| 5 | 1871 | 23 | 34 | 41 | 1 |
| 6 | 1498 | 24 | 28 | 42 | 1 |
| 7 | 1200 | 25 | 22 | 43 | 0.4 |
| 8 | 961 | 26 | 18 | 44 | 0.3 |
| 9 | 770 | 27 | 14 | 45 | 0.3 |
| 10 | 616 | 28 | 11 | 46 | 0.2 |
| 11 | 494 | 29 | 9 | 47 | 0.2 |
| 12 | 395 | 30 | 7 | 48 | 0.1 |

**Sources:** Author, Table (6) and equation (5).
\* For a community with an average population of 38.43 million and an average age of 34.96 years.

**Appendix (2):** Prediction table of COVID-19 cases per million and median age after 44.80 days of COVID-19 pandemic*

| M. age | Cases per 1M | M. age | Cases per 1M | M. age | Cases per 1M |
| --- | --- | --- | --- | --- | --- |
| 20 | 19 | 31 | 147 | 42 | 1149 |
| 21 | 23 | 32 | 177 | 43 | 1385 |
| 22 | 27 | 33 | 214 | 44 | 1669 |
| 23 | 33 | 34 | 258 | 45 | 2013 |
| 24 | 40 | 35 | 310 | 46 | 2426 |
| 25 | 48 | 36 | 374 | 47 | 2925 |
| 26 | 58 | 37 | 451 | 48 | 3526 |
| 27 | 70 | 38 | 544 | 49 | 4250 |
| 28 | 84 | 39 | 656 | 50 | 5124 |
| 29 | 101 | 40 | 790 | 51 | 6177 |
| 30 | 122 | 41 | 953 | 52 | 7446 |

**Sources:** Author, Tables (6) and equation (7).
\* For a community with an average population of 38.43 million and an average temperature of 12.34 °C.

## Footnotes

2 BBC News (2020), Coronavirus: **Italian economy takes a body blow**, available at: https://www.bbc.com/news/business-51650974, (Accessed: 3 April 2002)

3 European Commission, **Internal Market, Industry, Entrepreneurship and SMEs**, Available at: https://ec.europa.eu/growth/tools-databases/regional-innovation-monitor/base-profile/madrid, (Last accessed: 8 April, 2020)

4 South China Morning Post (2020), **Why Wuhan is so important to China’s economy and the potential impact of coronavirus**, Available at: https://www.scmp.com/economy/chinaeconomy/article/3047426/explained-why-wuhan-so-important-chinas-economy-and-potential, (Accessed: 7 April, 2020)

5 European Commission, **Internal Market, Industry, Entrepreneurship and SMEs**, Available at: https://ec.europa.eu/growth/tools-databases/regional-innovation-monitor/base-profile/bavaria, (Last accessed: 7 April, 2020)

